# Evaluating the Impact of Principal Component and Mixed Model Approaches on Polygenic Risk Score Portability to Diverse Ancestries in the UK Biobank

**DOI:** 10.64898/2026.08.17.26360388

**Authors:** Ajay Srinivasan Harikrishnan, Ciaran Michael Kelly

## Abstract

Polygenic risk scores (PRS) offer considerable potential for precision medicine. However, their predictive performance often attenuates when applied to populations that differ from the genome-wide association study (GWAS) training population. There are many potential sources of this portability problem, and one relatively under-explored contributor is the presence of residual confounding in GWAS summary statistics. In particular, confounding specific to the training population may contribute to predictive performance that does not transfer to other populations, such that improved control of population stratification could potentially improve PRS portability. Here, we investigated whether varying levels of population stratification adjustment, through the inclusion of principal components and the use of mixed models, altered PRS portability in three broad ancestry groups in the UK Biobank. The PRS were built using European training data for coronary artery disease and type 2 diabetes and subsequently evaluated in South Asian, African, and Latin American participants. We found that increasing PC adjustment did not produce a consistent trend in portability across ancestry groups or phenotypes, despite modest reductions in the LDSC intercept. However, substantial ancestry- and phenotype-specific effects on transferability were observed. Mixed-model association provided no significant change in PRS discrimination or portability. These findings highlight the need for a better understanding of the nature of residual confounding in PRS and whether improving the causal validity of GWAS results can ultimately improve the transferability of predictive accuracy between populations.

## Introduction

There is great potential in using the results of genome-wide association studies (GWAS) to build polygenic risk scores (PRS) for the prediction of certain common disorders [1]. PRS may be used as tools for population-level risk stratification and to allow for more efficient screening programmes and potentially low-risk interventions, such as statins or GLP-1 receptor agonists for coronary heart disease (CAD) and Type-2 Diabetes (T2D), respectively, in addition to lifestyle interventions for both [2, 3].

However, PRS do not generally perform as well in populations that differ from those included in the GWAS training population. This is known as the transferability or portability problem [4–6]. To achieve equitable advances in precision medicine, it is important to understand the sources of this attenuation in predictive performance, and considerable effort has been devoted to improving cross-ancestry prediction [7].

There are many factors which contribute to PRS performing poorly in ancestry groups that did not form the basis of the training population [6]. Firstly, linkage disequilibrium (LD) differences between populations can result in the association between top GWAS single nucleotide polymorphisms (SNPs) and any underlying causal variants being significantly weaker. Secondly, differences in allele frequencies between groups can affect the extent to which included SNPs actually explain a significant portion of trait variance. For example, if a risk allele is at a particularly low frequency in the target population, it will not contribute substantially to explaining phenotypic variance at the population-level, even with a moderate effect size.

A third source of potential attenuation is that the summary statistics generated during GWAS may suffer from both genetic and environmental confounding effects mediated by population structure. If the underlying confounding effects apply differently in other populations, then the performance of the resultant PRS models may differ substantially [8, 9]. Although this source of predictive attenuation is not thought to be as important in magnitude as LD and allele frequency differences, debate still remains around the extent to which confounding effects in GWAS have truly been eliminated and if they contribute to PRS performance issues [9, 10].

The two most popular approaches for minimizing the unwanted effects of confounding in a GWAS are the incorporation of principal components (PCs) of genetic variation as covariates during association and prediction, as well as the use of linear mixed models (LMMs), which account for distant genetic relatedness between individuals during association [11, 12].

However, there remains much debate on how to establish best practices for these methods. For instance, there have been various studies conducting GWAS and building PRS using UK Biobank (UKB) data, and they often include different numbers of PCs during training and deployment [13–15]. Likewise, there is conflicting evidence as to whether PC-based methods and LMM-based methods offer comparable benefits or are instead complementary in their effects [16, 17]. When confounding is found to be present in the summary statistics, reductions in the LD-score regression (LDSC) intercept can be taken as evidence that it has been reduced through population stratification adjustment methods [18].

Importantly, the relationship between reductions in confounding and PRS portability may not be straightforward. Confounding that is specific to the GWAS training population may contribute to predictive performance in that population without being transferable to other ancestry groups. In this case, reducing such confounding could be expected to improve portability by removing sources of prediction specific to the training population. Conversely, if confounding effects are shared and exploitable across populations, they may contribute to predictive performance in both the training and target populations. Removing these effects could therefore reduce predictive performance in the target population, even while representing an improvement in the causal validity of the PRS. Consequently, reductions in measures of confounding, such as the LDSC intercept, are not necessarily expected to correspond to consistent improvements in cross-ancestry portability.

Although much attention has been devoted to improving PRS portability by addressing allele frequency and LD differences, the extent to which best practices for accounting for confounding can change PRS performance in target populations of different ancestries has been relatively underexplored. Likewise, the relationship between portability and traditional measures of confounding such as the LDSC intercept are not well-understood. This study aimed to investigate how increasing the number of included PCs during association and prediction affected PRS performance across diverse ancestry groups. We also sought to assess whether trends in portability were correlated with reductions in the LDSC intercept and whether additional benefits in terms of portability could be gained through the use of mixed-model approaches. We hypothesized that there may be phenotype- and ancestry-dependent effects on PRS portability as the method of population structure adjustment was varied.

## Methods

### Study Approach

We used European GWAS summary statistics generated with varying levels of population stratification adjustment to build polygenic risk scores and evaluate their accuracy in three ancestry groups in the UKB: South Asian, African, and Latin American. We bootstrapped the calculation of the portability ratio for each PRS with reference to a European hold-out set for both CAD and T2D. We compared portability trends with changes in the LDSC intercept, a measure of confounding. We then ran a cluster-robust logistic model, which accounted for repeated measures on the same individuals, to assess whether the association between the PRS and disease status differed by ancestry and correction strategy. Finally, we investigated the added benefit of using mixed models in addition to PCA for the generation of GWAS summary statistics through a formal comparison of the accuracy of standard GWAS and mixed-model-based GWAS.

### Dataset and Ancestry Assignment

The UKB full genotype release data of 488,377 individuals were used for this analysis. Initial quality control filters were applied to the genotype data using a minor allele frequency threshold of 0.1% as well as SNP and individual missingness filters of 3%. SNPs in the HLA region and on the sex chromosomes were excluded due to the complexity of their analysis [19].

As previously described in Lello et al. 2020 and Kelly et al. 2026, individuals with ethnic background codes 1, 1001, 1002, or 1003 (self-reported White individuals) were selected as a European training population [20, 21]. An independent case-control hold-out set for each phenotype was set aside for PRS evaluation and portability calculation and was excluded from each GWAS described below. Cases and controls were matched in terms of the general age-gap differences observed between case-control pairs of siblings in a previous analysis using this data [21].

GRAFpop version 2.4 with 15,674 ancestry-informative SNPs was used to assign the non-White European UKB samples to specific population groups [22]. Three GRAFpop ancestry groups were deemed to have sufficient sample size for continued analysis: South Asian, African, and Latin American 1 (largely consistent with those of Puerto Rican and Dominican ancestry). Any individuals with a conflict between GRAFpop-assigned and self-reported ancestry were removed from the PRS analysis.

### GWAS and PRS

T2D and CAD were chosen as the phenotypes of interest due to their relatively large sample sizes among non-European ancestry individuals in the UKB and their promise in the clinical PRS arena. Final sample sizes of the case-control cohorts for each ancestry group across the two phenotypes can be seen in Table 1. UKB data field 22009 was used to access principal component information and a maximum of sixteen were used as per guidelines from previous research [14]. The training GWAS summary statistics were generated across varying levels of PC inclusion as previously described in Kelly et al. 2026 [21]. Briefly, for the PC-only association, PLINK2’s generalized linear model (GLM) was used, with the corresponding number of PCs included as covariates. For the linear mixed model-based GWAS, GCTA version 1.92.3 fastGWA-GLMM was used with genetic relatedness matrices (GRMs) constructed from 111,226 pruned SNPs [23, 24]. Summary statistics from both approaches were clumped with an LD threshold set to 0.1, a distance threshold of 500kb, and an index SNP significance threshold of 0.5. LDSC intercepts were also calculated for each set of summary statistics using LDSC software v.1.01.

**Table 1.** Baseline PRS performance and cross-ancestry portability.

| Phenotype | Ancestry | Cases | Controls | AUC | Portability Ratio |
| --- | --- | --- | --- | --- | --- |
| CAD | European | 3750 | 3752 | 0.567 | – |
|  | African | 315 | 3846 | 0.504 | 0.06 |
|  | South Asian | 1902 | 7550 | 0.546 | 0.70 |
|  | Latin American | 213 | 1498 | 0.503 | 0.06 |
| T2D | European | 2506 | 2510 | 0.631 | – |
|  | African | 773 | 3388 | 0.522 | 0.17 |
|  | South Asian | 2575 | 6877 | 0.561 | 0.47 |
|  | Latin American | 230 | 1479 | 0.607 | 0.81 |

The top 10,000 clumped SNPs were selected for each phenotype to build the PRS, based on performance in the training population and to facilitate direct comparisons between phenotypes. The PRS were constructed for each ancestry group separately using PLINK2. Each PRS was residualized against the corresponding number of PCs included in the training GWAS. In total, ten PRS (nine eight standard GWAS and an additional mixed-model GWAS), each with the same included top SNP-set, were calculated for each of the four ancestry groups.

### Bootstrapping and Statistical Analysis

Each PRS was evaluated on the case-control cohorts using the area under the receiver operating characteristic curve (AUC). Bootstrapping with 1,000 iterations was undertaken for AUC calculation to generate confidence intervals for predictive performance. The portability ratio was defined as the AUC on the target-ancestry population minus 0.5, divided by the AUC of the European hold-out set minus 0.5.

A cluster-robust logistic regression model was fitted to the non-European PRS data using R version 4.2.1: [Disease Status*∼* PRS *×* Ancestry Group *×* PC-Correction Level] PC-correction level was factorized in order to allow for non-linear relationships in the effects of increasing the number of included PCs. The individual was used as the unit of clustering to account for the fact that the same individuals were used across each PRS [25]. Sandwich correction was used to correct the standard errors of the model. For this analysis, PRS values were standardized within each phenotype *×*ancestry *×* PC-level group. Omnibus Wald tests were used to test the significance of the three-way interaction between PRS, ancestry group, and PC-correction level for each phenotype. P-values were adjusted using the Benjamini-Hochberg (BH) procedure.

To investigate the added benefit of mixed models as opposed to standard GLMs, DeLong’s test for correlated AUCs was performed for each phenotype and ancestry group using the full set of sixteen PCs included. P-values were also adjusted using the Benjamini-Hochberg (BH) procedure.

## Results

### Baseline Accuracy and Portability

In order to establish baseline PRS performance without any attempt to control for population stratification, we first calculated the AUC and portability ratios using the relevant ancestry groups. Here, the summary statistics and PRS were generated without using any principal components as covariates, and the results can be seen in Table 1.

For both traits, there was a clear decrease in performance when moving from the European hold-out set to the other three ancestry groups. The smallest portability ratio was seen for the African cohorts, aligning with previous literature [4, 6].

For CAD, the South Asian test set had the highest portability ratio (0.70), but both the African and Latin American test set PRS barely discriminated between cases and controls above random chance. In contrast, the Latin American cohort had the highest portability ratio for T2D (0.81), with a modestly discriminating AUC value of 0.61.

### Changes in Portability Across Increasing Levels of PC Inclusion

We next investigated whether attempts to control for population stratification could affect the portability of the PRS to the different ancestry cohorts. To do this, we generated GWAS summary statistics with increasing numbers of PCs included as covariates during association and PRS construction. We also inspected whether any trends in portability were aligned with changes in the LDSC intercept with increasing PC inclusion. The results for this analysis for CAD can be seen in Fig. 1.

**Figure 1.**
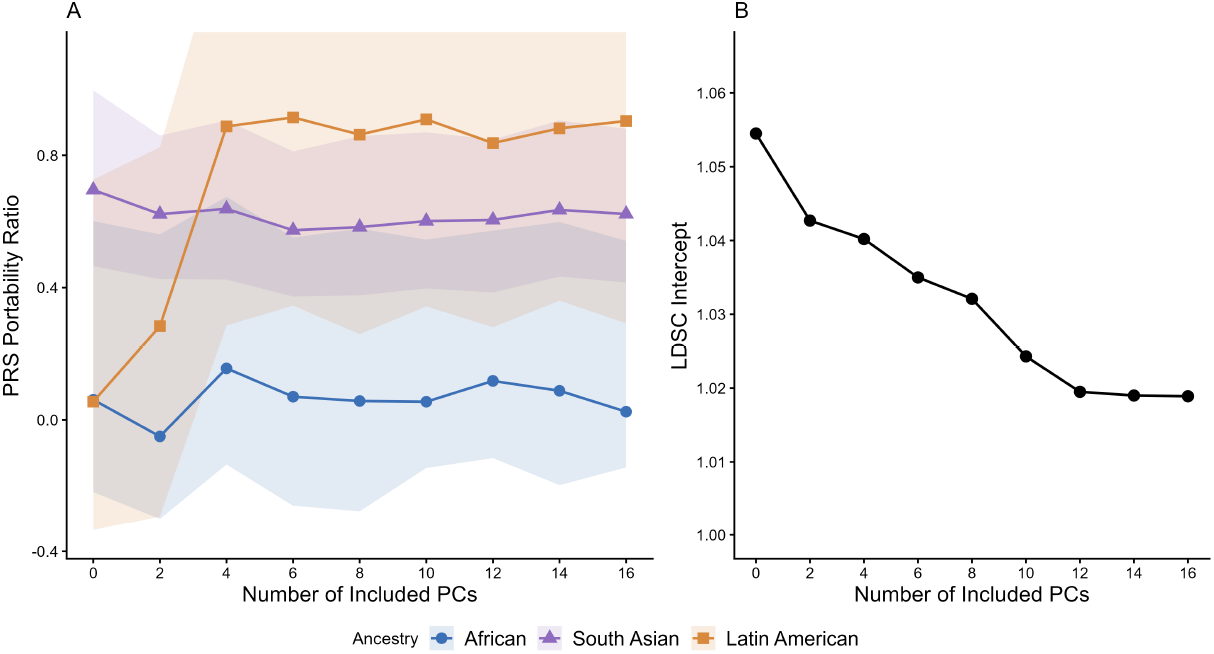
**(A)** Changes in CAD polygenic risk score portability across ancestry groups with increasing levels of principal component adjustment. Shaded regions represent 95% bootstrap confidence intervals. **(B)** Changes in the LDSC intercept derived from the European training population CAD GWAS summary statistics with increasing levels of principal component adjustment.

For the South Asian cohort, there was no strong effect of increasing the number of principal components on the portability of the PRS, despite modest reductions in the LDSC intercept.

In contrast, varying the number of included PCs appeared to have a clear effect on portability in the Latin American cohort. For CAD, portability increased substantially with the inclusion of 2 and 4 PCs, rising from close to zero to approximately 0.9. Although the bootstrap confidence intervals were wide, indicating considerable uncertainty around the precise portability estimates, the overall trend towards increased portability with PC adjustment was clearly visible. This improvement then plateaued between 4 and 16 PCs. The pattern broadly mirrored the decrease in the LDSC intercept up to 4 PCs. However, while portability showed little further improvement beyond 4 PCs, the LDSC intercept continued to decrease up to 12 PCs.

Lastly, the African CAD cohort showed greater variability in portability. No clear linear trend was observed, with fairly wide confidence intervals, and portability showing modest fluctuations depending on the number of included PCs.

The results for T2D can be seen in Fig. 2. Once again, for the South Asian cohort, there was no strong effect of increasing the number of principal components on the portability of the PRS, despite some small reductions in the LDSC intercept. For the PRS evaluated on the Latin American cohort, the effect of increasing PC inclusion up to four PCs was in the opposite direction to that observed for CAD, albeit less pronounced. Here, portability decreased modestly as more PCs were included in the model.

**Figure 2.**
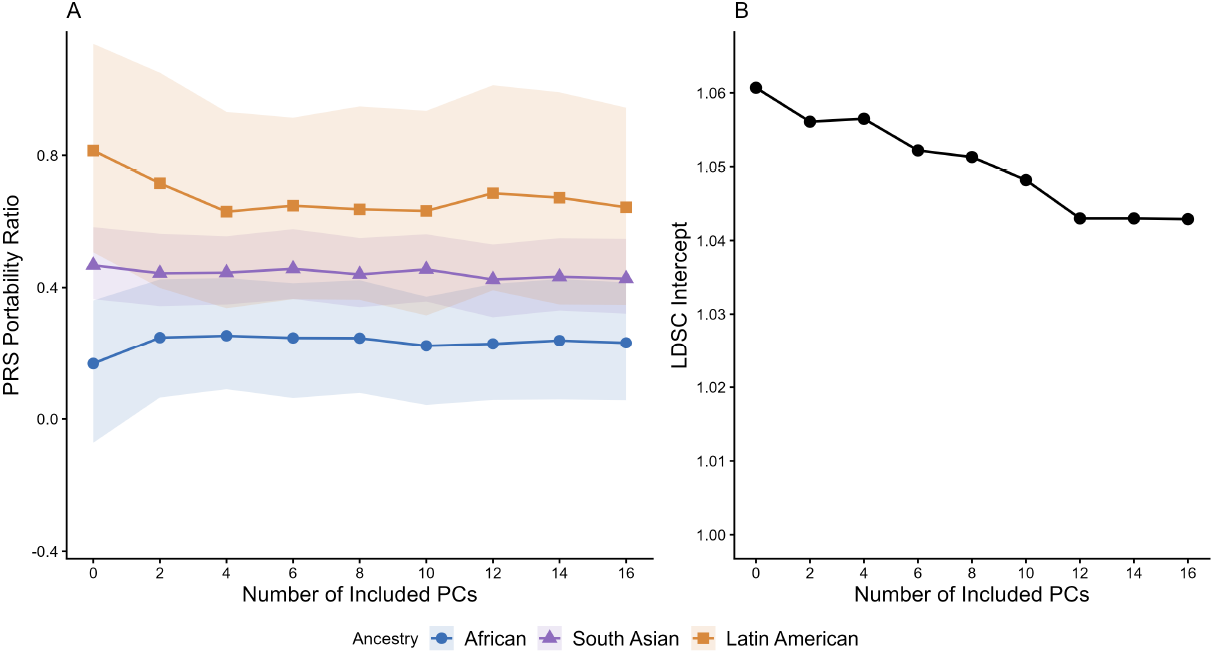
**(A)** Changes in T2D polygenic risk score portability across ancestry groups with increasing levels of principal component adjustment. Shaded regions represent 95% bootstrap confidence intervals. **(B)** Changes in the LDSC intercept derived from the European training population T2D GWAS summary statistics with increasing levels of principal component adjustment.

For the African T2D cohort, a small increase in portability was seen with the inclusion of the first two PCs. However, there was no subsequent noticeable change in portability with increasing PC inclusion.

### Disease Association With PRS, Ancestry, and PC Correction Level

We next investigated whether varying the number of PCs used changed the relationship between the PRS and disease differently across the three test ancestries. In order to do this while accounting for repeated observations of the same individuals across the various PC-corrected PRS analyses, we ran a logistic model with cluster-robust standard errors. The significance of the interaction terms between PRS, ancestry, and PC level for the two phenotypes can be seen in Table 2.

**Table 2.** Omnibus Wald tests of three-way interaction between polygenic risk score, ancestry group, and principal component correction level.

| Phenotype | $\chi^2$ | df | $p$ | $p_{\text{adj}}$ |
| --- | --- | --- | --- | --- |
| CAD | 36.4 | 24 | 0.051 | 0.051 |
| T2D | 39.6 | 24 | 0.024 | 0.048 |

For both phenotypes, the results provided limited evidence for a three-way interaction between PRS, ancestry, and PC-correction level. The association did not reach statistical significance for CAD and although was marginally significant for T2D after multiple-testing correction.

### Additional Effects of Mixed Model Association

Lastly, we investigated the potential added benefit of performing mixed-model association alongside PC correction during the generation of GWAS summary statistics. This was done as there is conflicting evidence in the literature as to whether or not the two strategies are complementary or redundant.

DeLong’s test for correlated AUCs was performed for the PRS generated using results from the GLM and LMM models, both utilizing the full sixteen principal components of variation as covariates. The results for this analysis can be seen in Table 3.

**Table 3.** DeLong’s test comparing polygenic risk score discrimination between models built using standard GLM-based GWAS and mixed-model-based GWAS. Both GWAS incorporated 16 principal components (PCs) as covariates during association analysis.

| Phenotype | Ancestry | AUC<sub>GLM</sub> | AUC<sub>LMM</sub> | $p$ | $p_{\text{adj}}$ |
| --- | --- | --- | --- | --- | --- |
| CAD | European | 0.574 | 0.574 | 0.626 | 0.715 |
|  | African | 0.502 | 0.505 | 0.022 | 0.181 |
|  | South Asian | 0.546 | 0.546 | 0.489 | 0.715 |
|  | Latin American | 0.567 | 0.565 | 0.092 | 0.369 |
| T2D | European | 0.633 | 0.633 | 0.715 | 0.715 |
|  | African | 0.531 | 0.530 | 0.589 | 0.715 |
|  | South Asian | 0.557 | 0.556 | 0.605 | 0.715 |
|  | Latin American | 0.585 | 0.586 | 0.562 | 0.715 |

We found no evidence that the LMM approach altered AUC or portability relative to the standard GLM approach.

## Discussion

PRS portability from one population to another is influenced by many factors, including differences in LD structure and allele frequencies. However, the focus of this research was on the potential contribution of confounding and population structure effects to performance loss. The aim of this research was to measure the extent to which increasing population stratification control changes the portability of PRS developed in European-ancestry individuals to non-European population groups in the UK Biobank. We did not find, as a general rule, that increasing the number of PCs included during training and prediction led to improved performance in the target population.

Importantly, across the two phenotypes examined, increasing the number of PCs did modestly lower the LDSC intercept, which is a traditional measure of confounding. However, across the three ancestry groups examined, there was no consistent pattern as to whether lowering the LDSC intercept would increase or decrease portability. For this reason, reductions in the LDSC intercept should not be taken to mean that portability is likely to be improved or diminished. Here, it is important to again note that the lack of improved performance as more PCs are added does not imply that population stratification control or confounding reduction was not ultimately successful. Indeed, if the confounding effects are shared and exploitable between populations, then removing them from the PRS model would reduce the overall predictive accuracy in a desirable manner. Without a better understanding of the underlying nature of the confounding (i.e., the specific environmental or genetic background effects being exploited), it will be hard to know if predictive portability would increase or decrease as confounding is reduced. Our findings highlight the complex relationship between confounding and PRS portability and demonstrate the need for further investigation into how the nature of confounding influences the transferability of polygenic predictions.

When looking at all the data combined in a mixed model, there was not compelling evidence for a specific effect of the association between PC correction level and the association between polygenic risk scores and the phenotype that differed by ancestry. This was significant for the T2D phenotype, but only barely so. For CAD, this effect was just beyond statistical significance. The general lack of effect on accuracy in either phenotype among the South Asian cohort could have contributed to this result, and although an ancestry-specific effect was not strongly supported by the model, it might be worthwhile for future studies to separate ancestry groups when looking at the disparate effects of population stratification control.

The most striking effect specific to an ancestry and phenotype was the performance of the T2D PRS in the Latin American cohort. When no attempt at population stratification control was made, there was almost no portability of the PRS to that cohort, and the model barely distinguished cases from controls at a level beyond random chance. However, with the inclusion of four principal components of variation, there was upwards of 80% retention of predictive power when moving from the European cohort to Latin Americans. It is difficult to fully ascertain the source of this effect, but one possible explanation is that that confounding present in the European subset of the UK Biobank is not exploitable in the Latin American cohort and that accounting for population structure removes this effect. However, no strong effect was seen in the South Asian cohort, and it is only moderately present in the African cohort, which raises questions as to the nature of the effect. Further study would be needed to see if this effect could be replicated in ancestry groups recruited independently of the UK Biobank initiative.

There has been debate regarding the additional benefit of mixed models in improving confounding control when including PCs during genetic association. At least in terms of portability, we did not find that the mixed-model approach offered any additional benefit or hindrance in terms of predictive performance.

Limitations of this analysis include the fact that only two disease phenotypes were deemed to have sufficient sample sizes and potential PRS predictive power in the non-European ancestry contingent of the UK Biobank to be worth investigating here. Broader trends across more diverse phenotypes could not be examined, and it would be worthwhile to investigate this issue with larger sample sizes and a much wider variety of phenotypes. Furthermore, the extent of confounding within the UK Biobank training population used, which predominantly comprises individuals of White British ancestry, may be relatively limited. Although moderate confounding in the traits was indicated by the LDSC intercept, the extent of this confounding was not particularly large even in the absence of adjustment for population structure. Larger and more diverse GWAS training cohorts might show stronger effects than those observed here.

We did not model the effects of additional phenotype-specific covariates that may be included in certain PRS models and may affect the results observed here. We chose to focus only on the use of PCs as covariates during training and prediction.

A final consideration to note is that the case-control cohort of the independent White hold-out set were selected to have an age-gap distribution similar to that of sibling pairs investigated in a previous analysis. However, the cohorts among the other ancestries were not age-matched in this way due to sample size limitations. This may affect the magnitude of the portabilities seen, but should not substantially affect the interpretation of the overall results or any trends observed.

In conclusion, the results presented here do not demonstrate that increasing levels of principal component adjustment improve cross-ancestry prediction in the UK Biobank. Additionally, there was little benefit to the addition of mixed models in terms of PRS portability. Decreases in the LDSC intercept did not consistently track with improvements in portability. This may reflect differences in the extent to which confounding is shared across populations. While removing confounding specific to the training population would be expected to improve portability, removing confounding that is shared across populations could instead reduce predictive performance in the target population. In some instances, strong phenotype- and ancestry-group-specific effects were observed, highlighting the need for further investigation in this area. The extent to which improving the causal validity of PRS might aid polygenic prediction portability should therefore continue to be explored.

## Data Availability

The relevant scripts and data outputs are available from https://github.com/ciaranoceallaigh96/portability

## Acknowledgments

This study was conducted using the UK Biobank Resource (Application Number 103770). We thank the participants of the UK Biobank for the use of their data.

## Supporting Information

The relevant scripts and data outputs are available from http://github.com/ciaranoceallaigh96/portability.

